# Genomic Sequencing Surveillance and Antigenic Site Mutations of Respiratory Syncytial Virus in Arizona, USA

**DOI:** 10.1101/2023.06.16.23291299

**Authors:** LaRinda A. Holland, Steven C. Holland, Matthew F. Smith, Victoria R. Leonard, Vel Murugan, Lora Nordstrom, Mary Mulrow, Raquel Salgado, Michael White, Efrem S. Lim

**Author notes:** Address for correspondence: Efrem S. Lim, Arizona State University, PO Box 876101, Tempe, AZ 85287, USA. LaRinda A. Holland and Steven C. Holland contributed equally to this work. Author order was determined by mutual agreement.

## Abstract

We conducted Respiratory Syncytial Virus (RSV) genomic sequencing surveillance of 100 RSV-A and 27 RSV-B specimens collected between November 2022 and April 2023 in Arizona, USA. We identified mutations in the pre-fusion F protein antigenic sites in both RSV-A and RSV-B. Continued genomic surveillance will be important to ensure RSV vaccine effectiveness.

## Research

Respiratory syncytial virus (RSV) is an RNA virus of the *Paramyxoviridae* family which causes acute respiratory infections primarily in children, adults with severe lung disease, and the elderly (1). The United States experienced an early surge in cases of RSV during the 2022-2023 respiratory pathogen season (2). The surge in RSV infections coincided with high circulating levels of influenza and SARS-CoV-2 viral infections, particularly in children (2). In Arizona, USA, laboratory-confirmed RSV cases increased from September 2022 through March 2023 with cases peaking in mid-November (**Figure 1A**).

**Figure 1.**
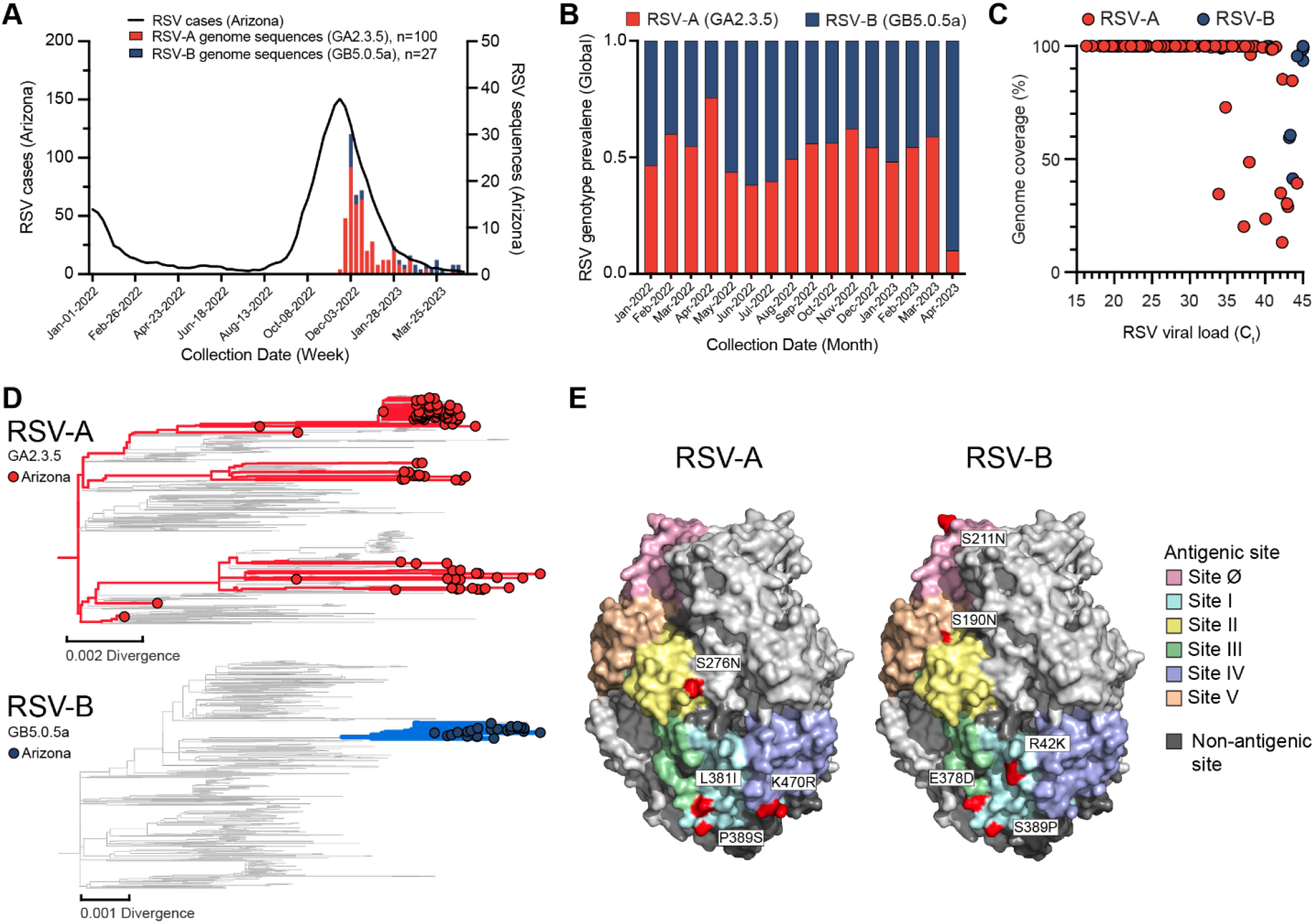
Genomic sequencing analysis of RSV in Arizona, USA, 2022-2023. (**A**) The 5-week moving average of PCR confirmed RSV detections in Arizona as reported by the National Respiratory and Enteric Virus Surveillance System (NREVSS) is shown alongside RSV sequence counts by genotype obtained for specimens utilized in this study. (**B**) The relative abundance of RSV-A and RSV-B genotypes is shown for all RSV genomes deposited in GISAID with collection dates between 1/1/2022 – 5/1/2023 (RSV-A n = 1,047; RSV-B n = 941) including genotypes obtained for specimens utilized in this study (RSV-A n = 100; RSV-B n = 27). (**C**) RSV RT-PCR Ct values and genome coverage for RSV-A (red) and RSV-B (blue) samples. (**D**) Phylogeny of the RSV-A GA2.3.5 (top) and RSV-B GB5.0.5a clades (bottom). Nodes and colored branches highlight Arizona sequences. (**E**) Structure of RSV prefusion F protein with surface exposed RSV-A (left) and RSV-B (right) SNPs found in Arizona sequences labeled and highlighted in bold red. One protomer of each trimer model is colored by antigenic site, two additional protomers shown in white.

The 2 major subtypes of RSV, RSV-A and RSV-B, have distinct antigenic differences in the P, N, F and G proteins (1). Each subtype is classified into genotypes based on sequence variability in the G protein (3). Based on sequencing data, the global distribution of RSV genotypes in 2022-2023 was split between the GA2.3.5 genotype of RSV-A and the GB5.0.5a genotype of RSV-B (**Figure 1B**). However, currently approved RSV vaccines are based on prefusion conformation of the F protein. Therefore, RSV genotypes do not reflect circulating RSV strains that harbor F protein mutations which can affect vaccine responses. Thus, genomic sequencing surveillance is needed to better understand the evolution of the virus and its potential impact on vaccine efficacy.

We performed genomic sequencing surveillance of RSV circulating in Arizona, USA, during the 2022-2023 season using remnant RSV-positive nasopharyngeal swabs (N=127) collected as part of standard-of-care respiratory pathogen testing at Valleywise Health (**Figure 1A**), which serves the population of Maricopa County. Next-generation sequencing (Illumina, 2 × 150 paired end) was performed using a hybrid-capture method targeted for the RSV genome (Illumina Respiratory Virus Oligo Panel v2). Sequencing reads were quality filtered, adapter trimmed (Trim Galore version 0.6.10), mapped to RSV-A and RSV-B reference sequences (GISAID EPI_ISL_412866 and EPI_ISL_165399) (Burrows-Wheeler Aligner version 0.7.17-r1188), and consensus sequences generated (SAMtools version 1.17). We assembled the complete genome sequences of 92 RSV-A (GA2.3.5 genotype) and 24 RSV-B (GB5.0.5a genotype), and partial genome sequences of 8 RSV-A (GA2.3.5) and 3 RSV-B (GB5.0.5a) (GenBank Accessions: OR143134 - OR143250; GISAID Accessions: EPI_ISL_17808760 - EPI_ISL_17808814). To determine RSV viral load, we performed qRT-PCR assays (HRSV-pan) that recognizes both subtypes RSV-A and RSV-B (4). The mean RSV CT value was 29.83 with a standard deviation of 7.44. We found that specimens with viral load CT’s ≤ 33 yielded 99-100% genome coverage (**Figure 1C**). Whole genome phylogenetic analysis (Nextclade version 2.14.1) showed that the Arizona RSV-A sequences were polyphyletic in the GA2.3.5 clade indicating at least 3 independent introductions of RSV-A into Arizona (**Figure 1D**). The Arizona RSV-B sequences formed a monophyletic clade in GB5.0.5a indicating a single introduction that seeded local transmission within the state (**Figure 1D**). Our findings are consistent with investigations of RSV in Massachusetts (5) and Washington (6) that infer that the atypical increase in cases during the 2022-2023 season was the result of multiple introductions of extant lineages and not attributable to a divergent RSV lineage with increased virulence or transmissibility.

To date, two RSV vaccines (Arexvy and Abrysvo) have been FDA approved for individuals 60 years and older in the US. Both vaccines are based on the RSV prefusion F protein with Arexvy being monovalent and Abrysvo bivalent (7, 8). The majority of host antibodies target six antigenic sites of the F protein, characterized as sites Ø-V (9). We examined the F gene sequences of the Arizona RSV genomes and identified 7 non-synonymous substitutions in antigenic sites I, II, IV, and V of RSV-A (**Table 1**). Similarly, we identified 5 non-synonymous substitutions in the antigenic sites Ø, I, II, and V of RSV-B. All mutations in RSV-A genomes were found in low frequency, with no mutations observed in greater than 9% of samples. Conversely, in the RSV-B genomes, most presented with high frequency mutations and only one rare SNP was found. These trends in mutation frequencies were comparable to the frequency in global RSV genome sequences. Finally, we mapped the mutations onto the pre-fusion F protein crystal structure (PDB: 7KQD). The structural model overlay revealed that the mutations are exposed on the F protein surface suggesting that they may interfere with antibody binding (**Figure 1E**).

**Table 1:** Non-synonymous amino acid substitutions in RSV-A and RSV-B F protein antigenic sites found in Arizona genome sequences compared to global genome sequences.

| RSV-A | Mutation | Antigenic site | Arizona frequency (n=92) | Global frequency (n=952) |
| --- | --- | --- | --- | --- |
|  | I57V | Site V | 2 (2%) | 20 (2%) |
|  | I59V | Site V | 1 (1%) | 1 (0%) |
|  | S276N | Site II | 4 (4%) | 162 (17%) |
|  | V379A | Site I | 8 (9%) | 2 (0%) |
|  | L381I | Site I | 1 (1%) | 0 (0%) |
|  | P389S | Site I | 2 (2%) | 4 (0%) |
|  | K470R | Site IV | 1 (1%) | 0 (0%) |
| RSV-B | Mutation | Antigenic site | Arizona frequency (n=24) | Global frequency (n=894) |
|  | R42K | Site I | 18 (75%) | 95 (11%) |
|  | S190N | Site V | 24 (100%) | 555 (77%) |
|  | S211N | Site Ø | 24 (100%) | 554 (76%) |
|  | E378D | Site III | 2 (8%) | 0 (0%) |
|  | S389P | Site I | 24 (100%) | 566 (78%) |

Although RSV remains a significant clinical burden, the recently approved RSV vaccines reduce the risk of lower respiratory tract illness. By tracking the evolution of RSV, we can improve the design of vaccine formulations to improve vaccine effectiveness. A limitation of this study is that we do not yet understand the functional consequences of the mutations identified in the F protein antigenic sites. Nonetheless, our surveillance revealed sequence diversity in the F protein antigenic sites that is not reflected in current genotyping schema, which is based on G protein sequences. Overall, this study demonstrates the importance of genomic sequencing surveillance for RSV and other pathogens of interest to clinical and public health.

## Data Availability

All data produced in the present study are available upon reasonable request to the authors. RSV genome sequences have been deposited into the GenBank database under accession numbers OR143134 - OR143250; and the GISAID EpiRSV database under accession numbers EPI_ISL_17808760 - EPI_ISL_17808814.

## Acknowledgements

We gratefully acknowledge Sarah Namdarian for assistance with the collection of the clinical specimens, Alexis Thomas, Gabrielle Hernandez Barrera, Michelle Tan for assisting in specimen processing, and Regan Sullins for assisting with library construction. We thank the authors from originating laboratories responsible for obtaining the specimens and the submitting laboratories where genetic sequence data were generated and shared via the GISAID initiative and NCBI GenBank.

This study was approved by the Arizona State University Institutional Review Board (STUDY00011967) and was supported in part by Arizona State University, and the Centers for Disease Control and Prevention (CDC BAA 75D30121C11084).

## Data availability

RSV genome sequences have been deposited into the GenBank database under accession numbers OR143134 - OR143250; and the GISAID EpiRSV database under accession numbers EPI_ISL_17808760 - EPI_ISL_17808814.

## Competing interests

The authors declare no competing interests.

## Contributions

Conceptualization: E.S.L.; Formal analysis: L.A.H., S.C.H., M.F.S., V.R.L.; Investigation: L.A.H., S.C.H., M.F.S., V.R.L., E.S.L.; Resources: V.M., L.N., M.M., R.S., M.W.; Data curation: L.A.H., S.C.H., M.F.S., V.R.L.; Writing-original draft: L.A.H., S.C.H., M.F.S., E.S.L.; Writing-review and editing: L.A.H., S.C.H., M.F.S., L.N., E.S.L.; Supervision: E.S.L.; Funding acquisition: E.S.L. All authors reviewed and approved the final manuscript.

## Notes

### Competing Interest Statement

The authors have declared no competing interest.

### Funding Statement

This study was supported in part by Arizona State University, and the Centers for Disease Control and Prevention (CDC BAA 75D30121C11084).

### Author Declarations

This study was approved by the Arizona State University Institutional Review Board.

